# Effectivity and Safety Profile of Tenapanor, a Sodium-Hydrogen Exchanger Isoform 3 Inhibitor, as an Innovative Treatment for Hyperphosphatemia in Chronic Kidney Disease, a Systematic Review of Clinical Studies

**DOI:** 10.1101/2023.12.19.23300205

**Authors:** William Suciangto, Haerani Rasyid, Anastasya Angelica Vicente, Winny Suciangto

**Affiliations:** Faculty of Medicine, Hasanuddin University, Makassar City, South Sulawesi Province, Indonesia; Division of Nephrology and Hypertension, Department of Internal Medicine, Faculty of Medicine, Hasanuddin University, Makassar City, South Sulawesi Province, Indonesia

**Keywords:** Chronic Kidney Disease, Hyperphosphatemia, Tenapanor

## Abstract

**Background:** Chronic kidney disease (CKD) is a major global health problem. Hyperphosphatemia is frequent in CKD and a reason for increased morbidity and mortality as it generates hyperparathyroidism, high fibroblast growth factor 23 (FGF23), and hypocalcemia. Available hyperphosphatemia therapies still have limitations, including risk of metal overload, cardiovascular calcification, and systemic adverse effects (AEs). Tenapanor is a new hyperphosphatemia treatment in CKD with sodium-hydrogen exchanger isoform 3 (NHE3) inhibition mechanism and low systemic AEs.

**Objectives:** Discovering the effectivity and safety of tenapanor as hyperphosphatemia management in CKD.

**Method:** Literature searching is performed by using “pubmed” and “science direct” with “tenapanor”, “chronic kidney disease”, and “hyperphosphatemia” as keywords. The literatures were selected using PRISMA algorithm version 2020. Literature was screened based on Population, Intervention, Comparison, and Outcome (PICO) criteria which are: CKD patients requiring dialysis as population, tenapanor or its combination with dialysis or phosphate binders as intervention, placebo or other phosphate binders without tenapanor as comparison, and serum phosphate, safety profile, and other pleiotropic benefits related to hyperphosphatemia management as the outcome. The included studies then assessed for risk of bias and qualitatively reviewed.

**Outcome:** Tenapanor was able to reduce serum phosphate, generally in a dose-dependent manner. Tenapanor also suppressed FGF23 and parathyroid hormone, probably due to decreased serum phosphate. The frequent AEs was transient mild-to-moderate diarrhea in a dose-dependent manner. Tenapanor was generally well-tolerated with low systemic AEs due to its non-calcium, metal-free, and low-absorbed properties.

**Conclusion:** Tenapanor is an effective and safe option for hyperphosphatemia management in CKD.

## INTRODUCTION

Chronic kidney disease (CKD) is still a major global health problem, with approximately 697.6 million people worldwide were reported to have CKD in 2017, of which 1.2 million people died due to CKD.^1^ Hyperphosphatemia is often found in CKD. Declined renal function, particularly in advanced stage will lead to serum phosphate accumulation due to inability in excreting serum phosphate which is mostly originated from daily intestinal absorption. Osteocytes then release fibroblast growth factor 23 (FGF23) in order to promote renal phosphate excretion. Unfortunately, FGF23 also impair renal 1.25-Dihydroxycalcitriol production which disturb intestinal calcium absorption, resulting in hypocalcemia. Parathyroid glands then release parathyroid hormone (PTH) as a compensatory mechanism to increase serum calcium through bone resorption. Calcium and phosphate release from destructed bone will lead to cardiovascular calcification and bone disorders, thus increasing morbidity and mortality in patients with CKD. ^2,3^

Dialysis, phosphate binders, and calcimimetic agents are available modalities for hyperphosphatemia management in CKD, but they still have some disadvantages including flexibility limitation, hypocalcemia risk, as well as metal overload and cardiovascular calcification.^4–6^ In this review, we provide clinical evidences of the effectivity and safety profile of tenapanor in managing hyperphosphatemia in CKD through its sodium-hydrogen exchanger isoform 3 (NHE3) inhibition.

## METHOD

Literature searching was performed in Pubmed and Science Direct by using “tenapanor”, “chronic kidney disease”, and “hyperphosphatemia” as keywords. The studies then systematically selected by using The Preferred Reporting Items for Systematic Reviews and Meta-Analyses (PRISMA) algorithm (Figure. 1). Duplicate literatures were removed. Review articles, books, conferences abstracts, editorials, commentaries, letter to editor, and guidelines were excluded, while research studies were retrieved. Literature screening was performed by using Population, Intervention, Comparison, and Outcome (PICO) criteria which are: CKD patients requiring dialysis as population, tenapanor or its combination with dialysis or phosphate binders as intervention, placebo or other phosphate binders without tenapanor as comparison, as well as serum phosphate and safety profile as the outcome. Studies which meet the criteria then examined for risk of bias as quality assessment and qualitatively synthesized to establish this systematic review.

**Figure 1.**
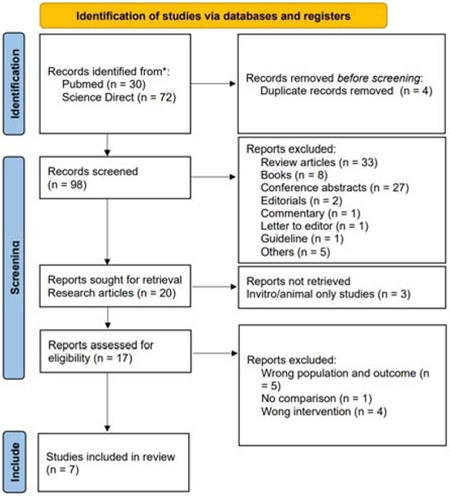
Literature Screening Algorithm by Using “The Preferred Reporting Items for Systematic Reviews and Meta-Analyses” (PRISMA) version 2020.

## RESULT AND DISCUSSION

### Risk of Bias Assessment in Included Studies

Our included studies have a relatively similarities in risk of bias. The participants and investigators were all blinded in all of our included studies.^7–13^ There are two studies with unclear risk of randomization sequence method as it was not mentioned in the studies.^8,10^ Five of included studies have unclear risk of concealment of randomization allocation. ^8,10–13^ All of our included studies have a low risk of bias in completeness of reported outcomes.^7–13^ The general summary of the risk of bias in our included studies can be seen in figure 2.

**Figure 2.**
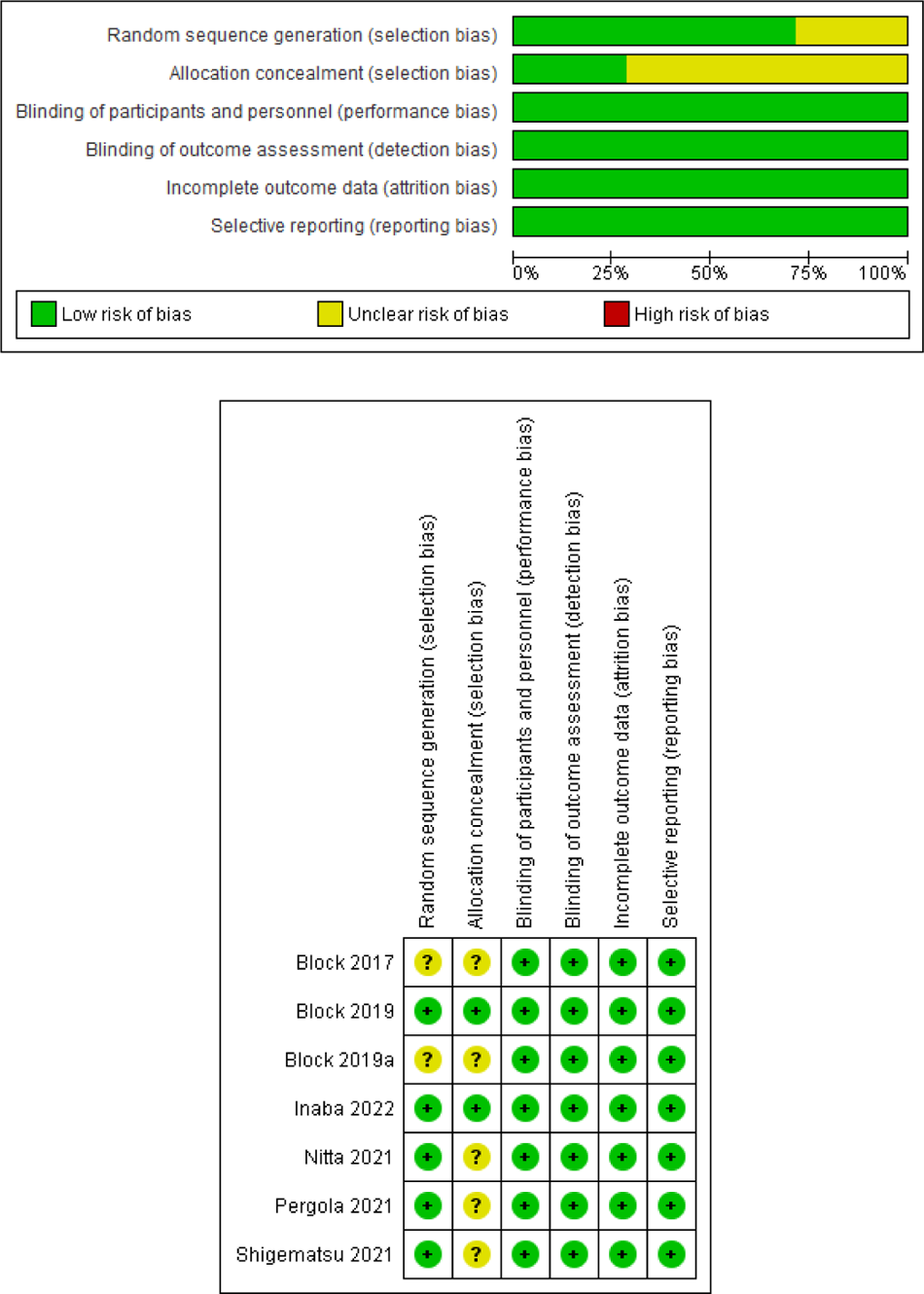
Risk of Bias Assessment of Included Studies.

### Characteristics of Included Studies

A total of seven clinical studies were included in our review. All the studies evaluated the decrease of serum phosphate as their primary outcome.^7–13^ Four studies use tenapanor or placebo alone as the intervention and comparison by implementing “washout system”^7–10^, while the three other studies still maintained the previous phosphate binders as a combination with tenapanor or placebo.^11–13^ Six of seven studies also examined FGF23 levels as the other outcome.^8–11,13,14^ The level of PTH was investigated in two studies.^9,13^ All of our included studies evaluated the safety profile of tenapanor. The summary of our included studies is described in table 1.

**Table 1.**
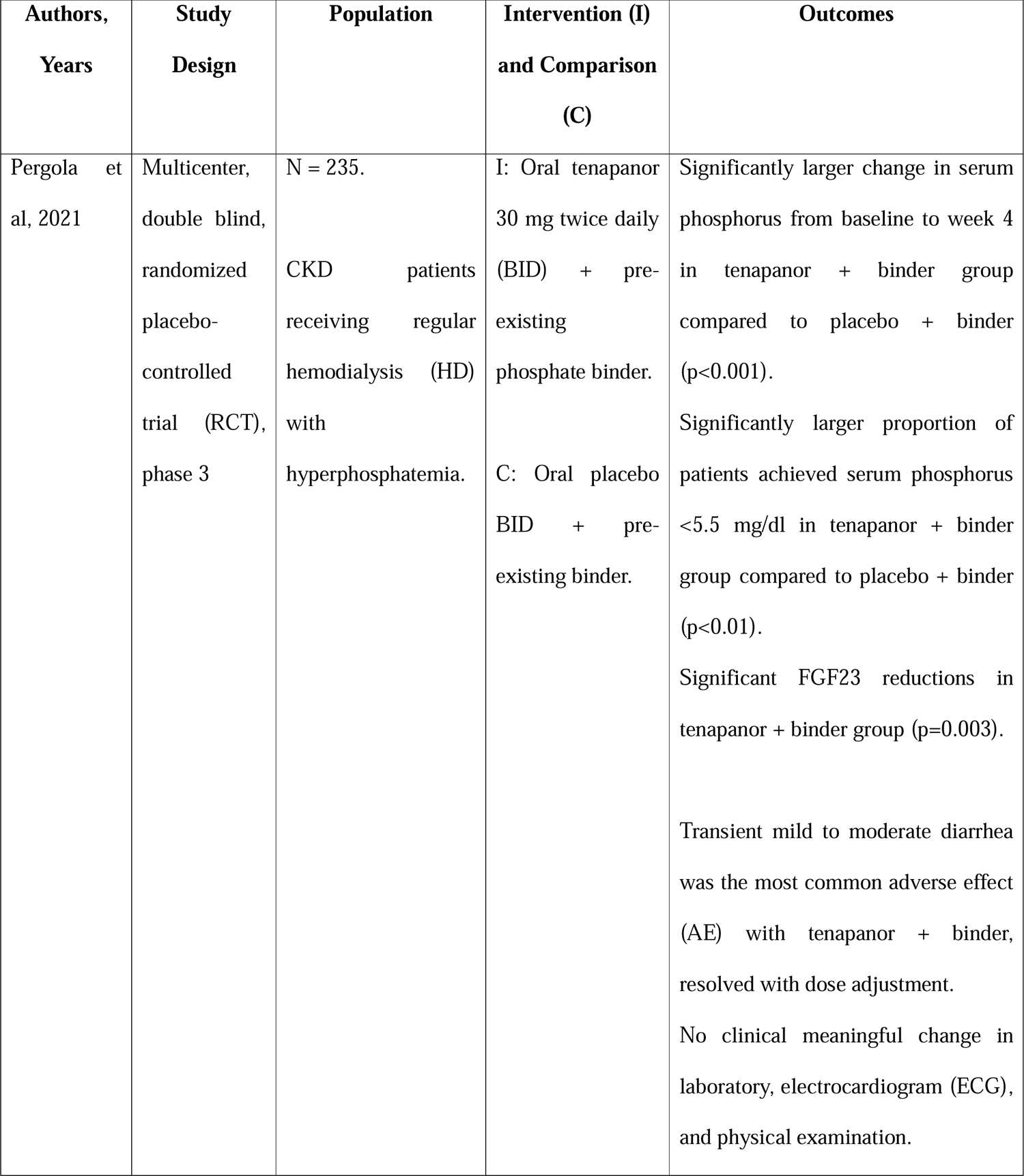
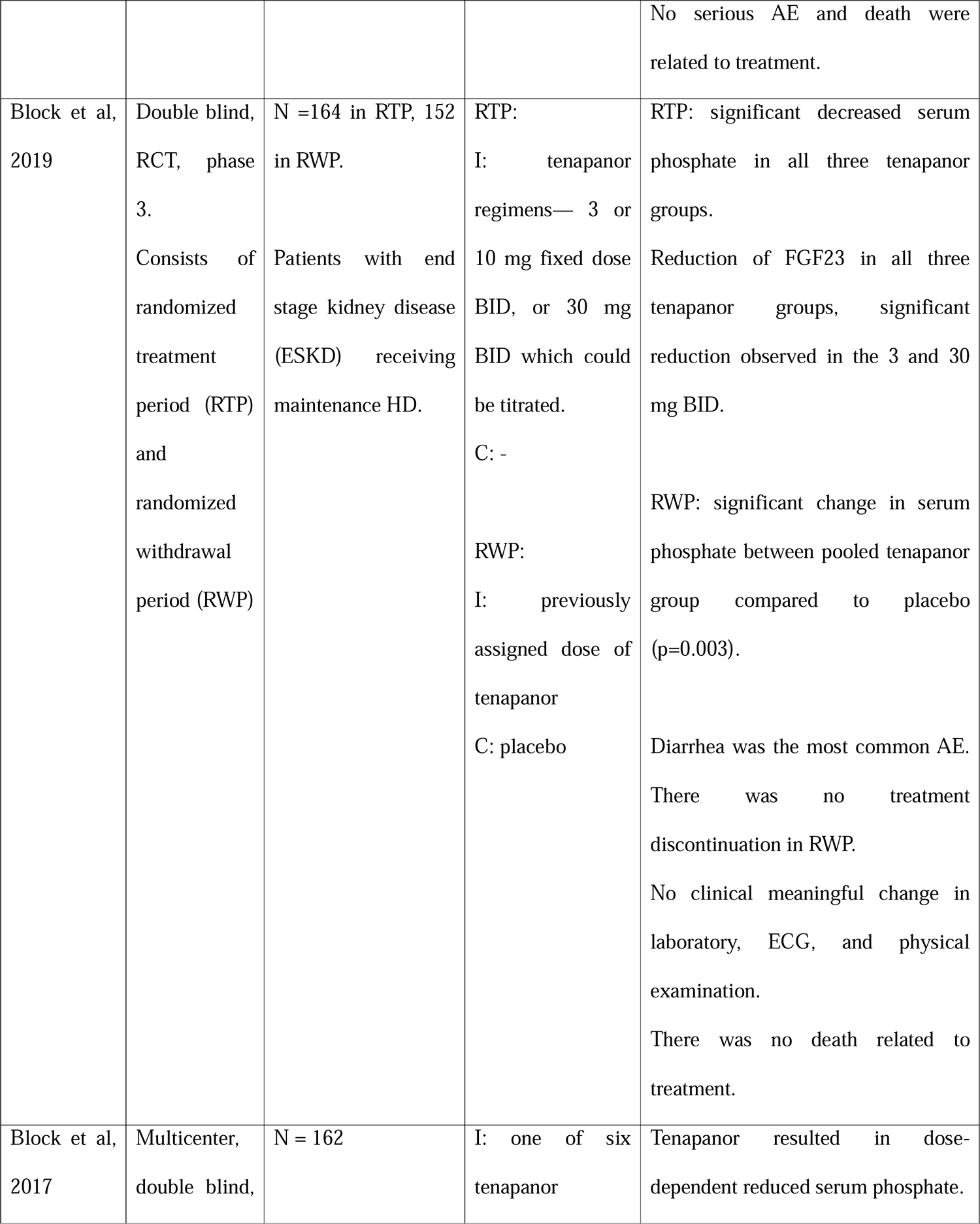
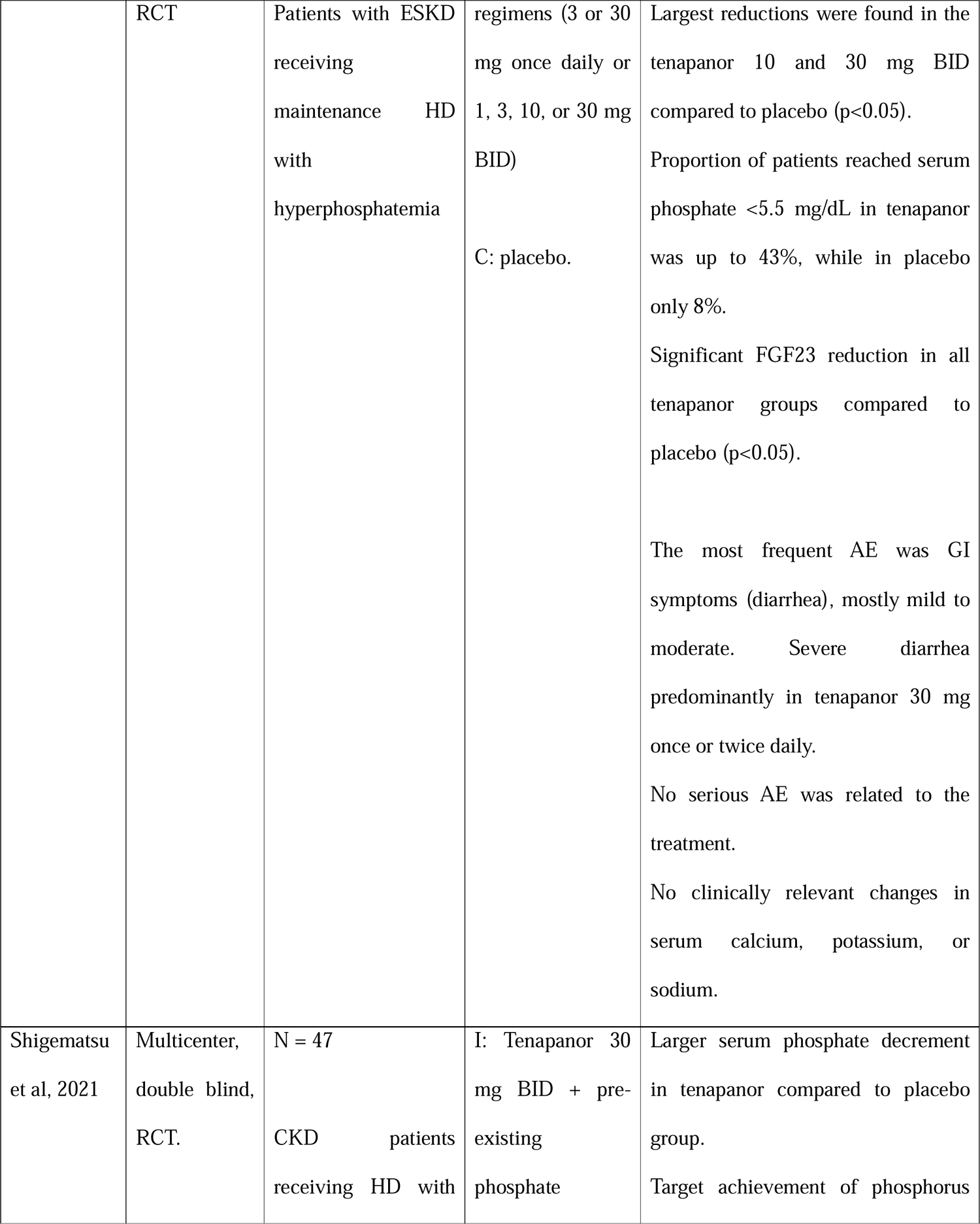
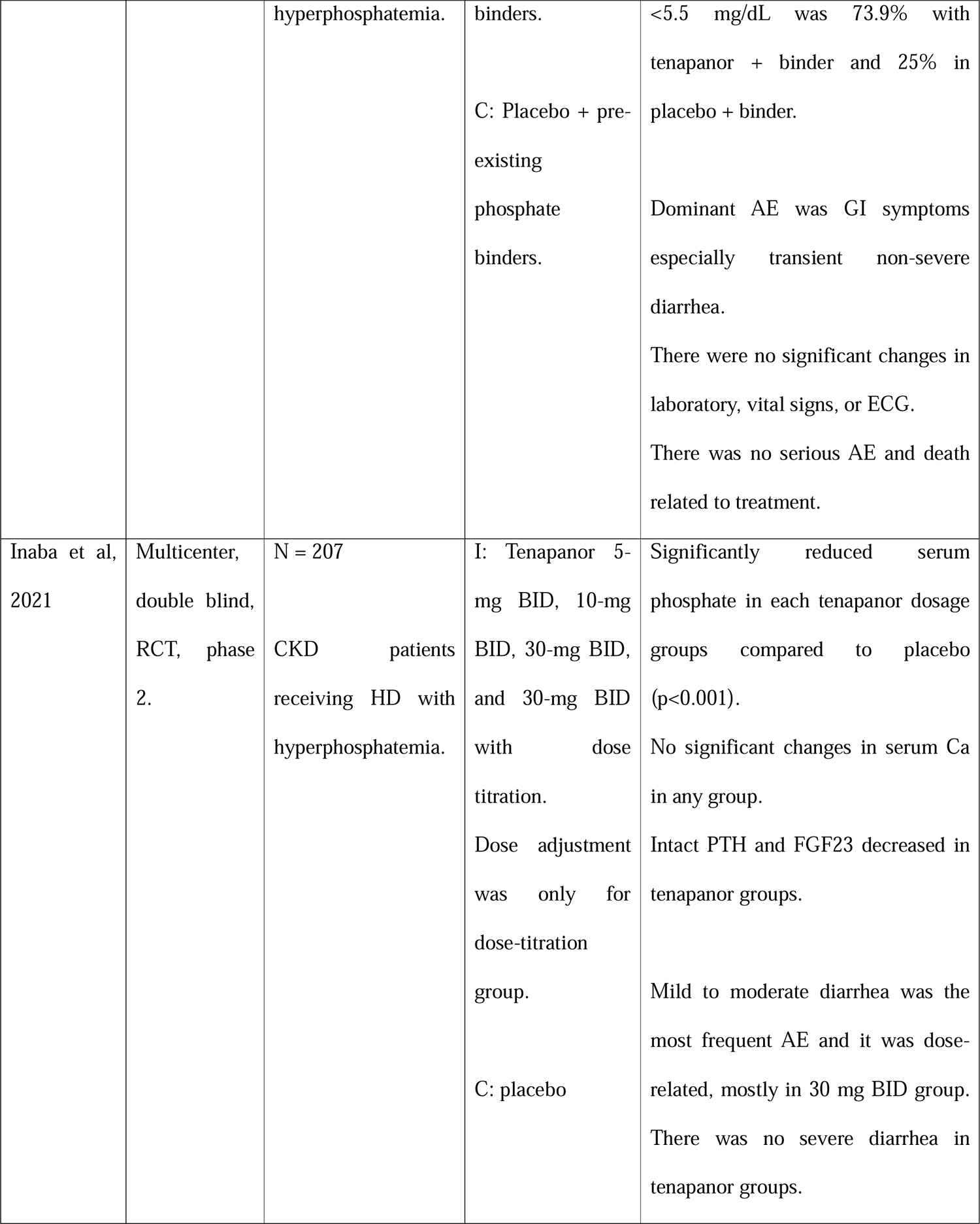
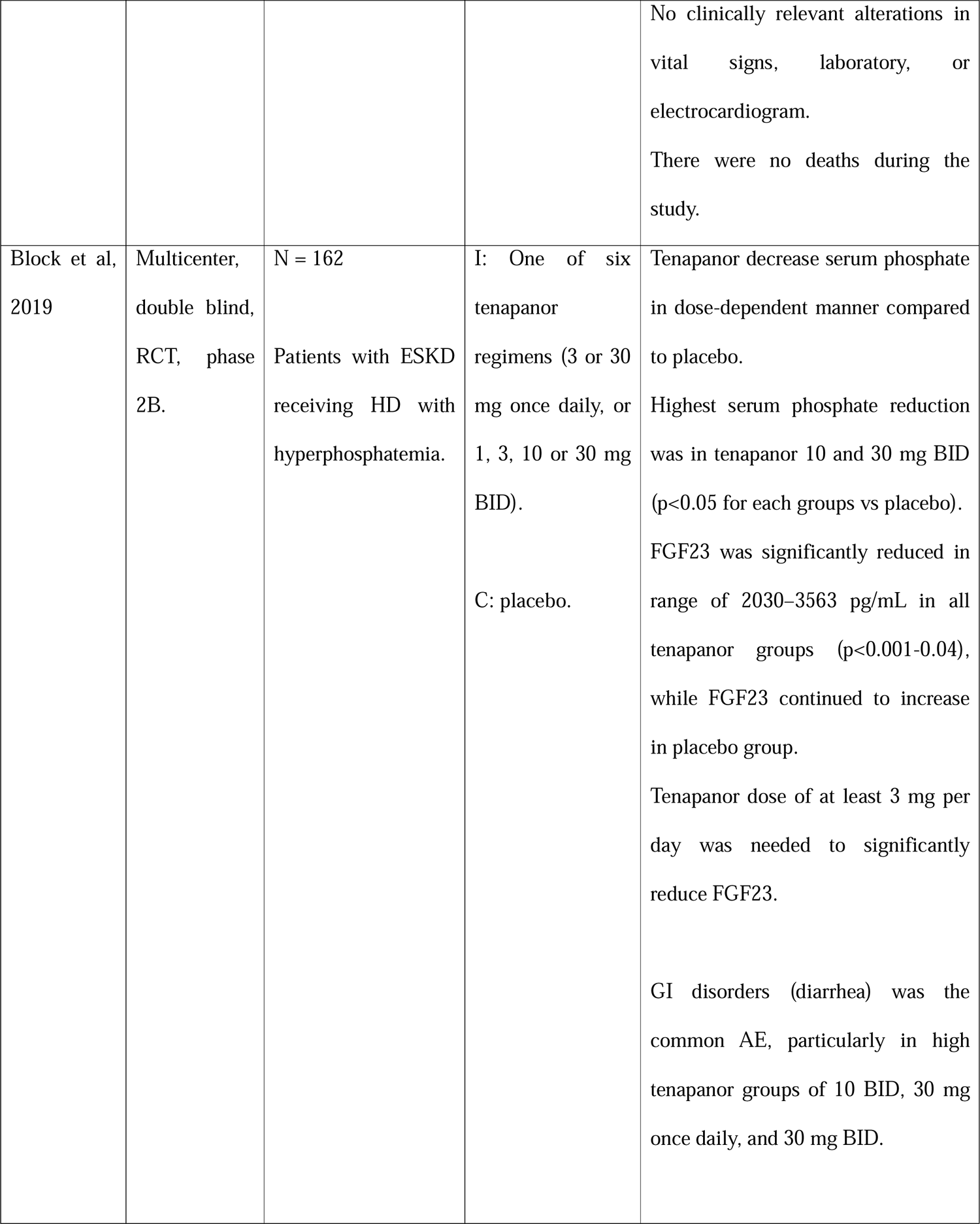
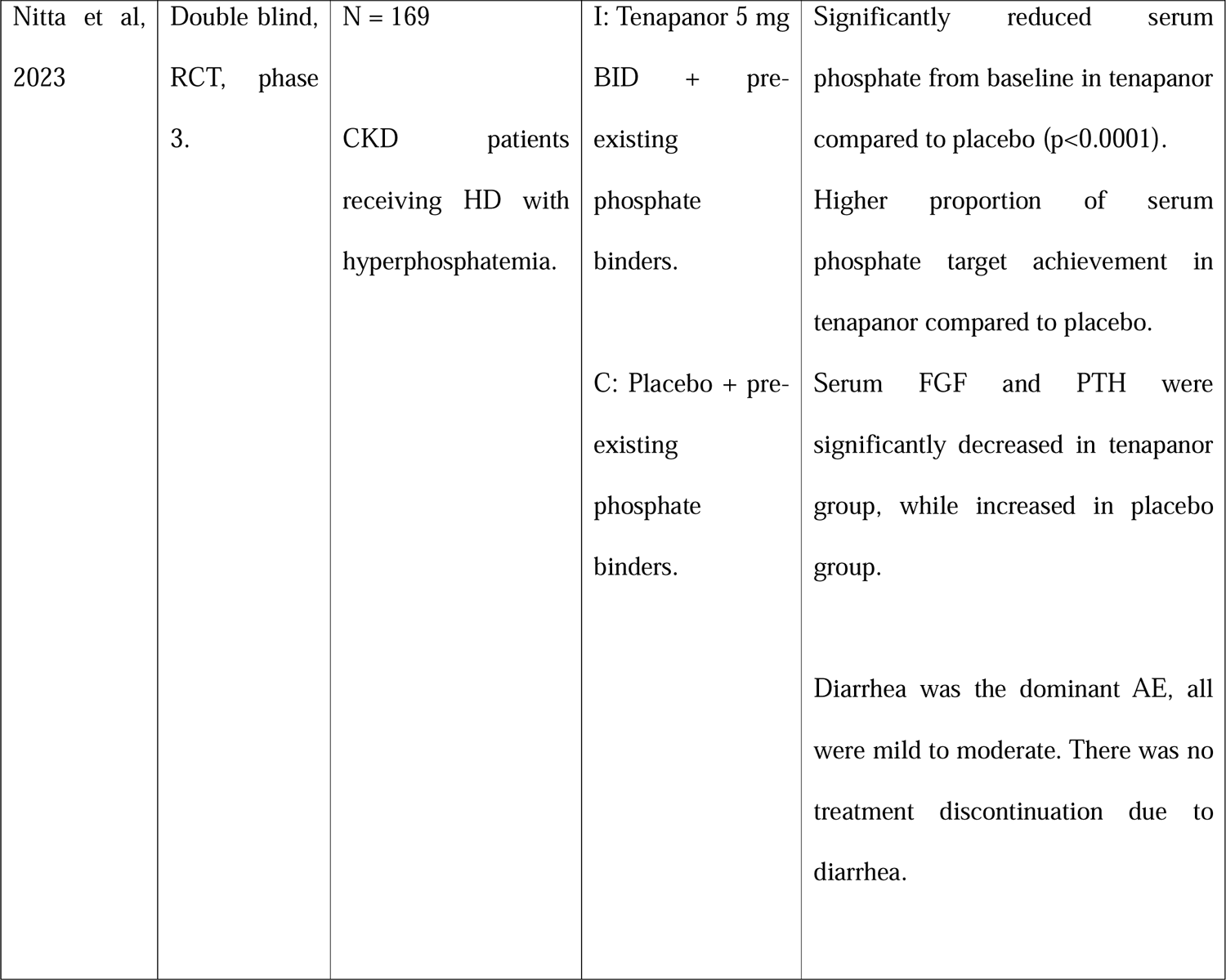
Table of Included Studies.

### The Role of Tenapanor as Hyperphosphatemia Treatment in Chronic Kidney Disease

Hyperphosphatemia is a frequent metabolic complication of CKD, particularly in end stage disease. This complication is a result of impaired renal function and limitation of conventional dialysis to eliminate serum phosphate originated from dietary phosphate absorption.^7,10^

Tenapanor is a selective inhibitor of the sodium-hydrogen exchanger isoform 3 (NHE3), an antiporter located on the surface of the gastrointestinal tract’s enterocytes. By inhibiting the sodium-hydrogen exchange, tenapanor causes the accumulation of intracellular protons and subsequently lowers the pH. This decrease in pH alters the tight junction proteins, resulting in a decrease in the permeability of phosphate paracellular diffusion and ultimately lowering the level of serum phosphate.^7–9,12^ The mechanism of tenapanor in inhibiting phosphate absorption can be seen in figure 3.

**Figure 3.**
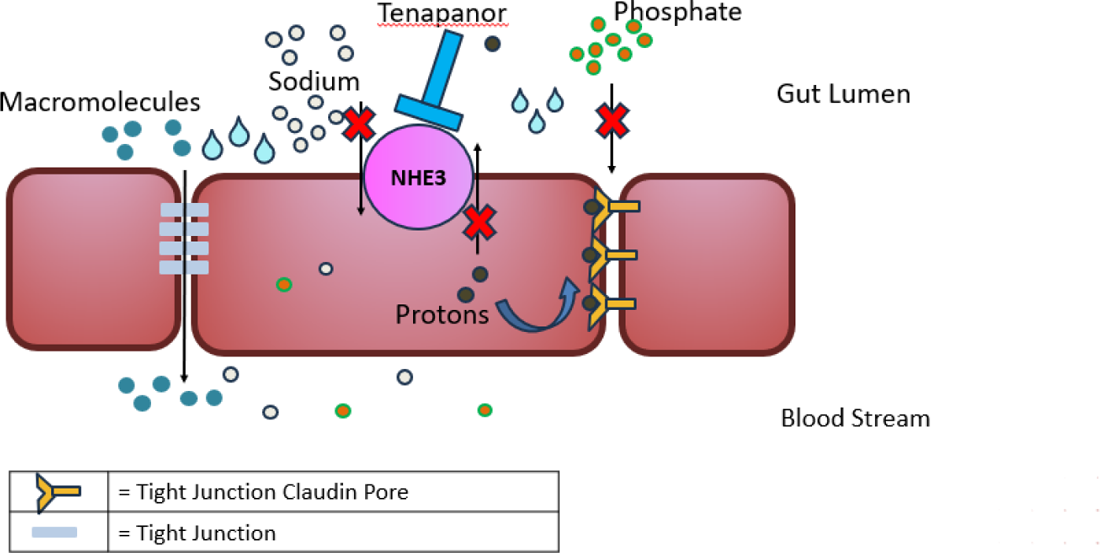
Tenapanor, Mechanism of Action

### Effectivity of Tenapanor in Lowering Serum Phosphate in Chronic Kidney Disease

There are four studies which investigated the effect of tenapanor as a single therapy to serum phosphate in CKD patients. All the patients underwent 1-3 weeks of washout period before started the treatment.^7–10^ The majority dose of tenapanor was 3, 10, and 30 mg twice daily (BID)^7–10^, though dosage of 3 and 30 mg once daily^8,10^ as well as 1 and 5 mg BID was also found^8–10^. All these four studies showed a higher decrease in serum phosphate with tenapanor as single therapy compared to placebo.^7–10^ Significant serum phosphate decrement from baseline was reported with tenapanor dose of 3, 5, 10, and 30 mg BID in fixed dose as well as 30 mg BID with titration versus placebo in two studies.^7–9^ Tenapanor reduces serum phosphate levels in a dose-dependent manner in all four studies.^7–10^ In the other hand, tenapanor as single therapy also resulted in higher proportion of patients achieving serum phosphate target of <5.5 mg/dL compared to placebo.^8^

Efficacy of tenapanor as combination therapy with other phosphate binders was observed in three studies. All the subjects in the studies maintained their pre-existing phosphate binders which then added with tenapanor or placebo.^11–13^ Two of three studies use tenapanor 30 mg BID as the treatment^11^, while one study use tenapanor 5 mg BID^13^. In all three studies, combination of tenapanor with phosphate binders revealed a greater reduction of serum phosphate compared to combination of placebo with phosphate binders.^11–13^ Statistical significance was found in two of three studies with tenapanor dose of both 5 mg BID and 30 mg BID.^11,13^ A higher rate of target achievement for serum phosphate was also found in tenapanor group compared to placebo in all three studies^11–13^, in which two of three studies set the serum phosphate target to <5.5 mg/dL^11,12^, while serum phosphate goal ranged from 3.5-6.0 mg/dL in one other study^13^.

### Pleiotropic Effect of Tenapanor in Interfering Fibroblast Growth Factor 23 and Parathyroid Hormone in Chronic Kidney Disease

Besides reducing serum phosphate, tenapanor was also found to have a beneficial effect on FGF23 and PTH levels. A total of six studies showed FGF23 declined with tenapanor^8–11,13,14^. Significant decrease in FGF23 was observed in tenapanor as single therapy at dose of 1, 3, 10, and 30 mg BID as well as 3 and 30 mg once daily.^7,8,10^ Tenapanor as combination therapy with other phosphate binders also showed its ability in reducing FGF23 levels at dose of both 5 mg and 30 mg BID.^11,13^

Tenapanor was also found to suppress PTH levels other than serum phosphate and FGF23 in two studies.^9,13^ As monotherapy, tenapanor at dose of 5, 10, and 30 mg BID was able to reduce PTH levels though the significance was not reported.^9^ In a setting as combination therapy with other phosphate binders, tenapanor 5 mg BID was reported to significantly suppress PTH.^13^

### Safety Profile of Tenapanor in Chronic Kidney Disease Population

Safety profile of tenapanor was evaluated in all of our included studies.^7–13^ Gastrointestinal symptoms especially diarrhea was the most frequent adverse effect (AE) in all the studies.^7–13^ The intensity of diarrhea was dominantly mild to moderate in severity with transient onset in majority of studies.^11–13^ In the setting of tenapanor as single therapy, the diarrhea tended to be dose-dependent, as the incidence of diarrhea was higher in tenapanor dose of 10 mg and 30 mg BID as well as 30 mg once daily.^7–10^ The same trend was also found in the setting of tenapanor as combination therapy with phosphate binders, in which higher incidence of diarrhea was reported in studies with higher dose of tenapanor.^11–13^ Severe diarrhea was reported in one study, which is predominantly in tenapanor dose of 30 mg once and twice daily.^15^

The safety profile of tenapanor was also evaluated with physical examination, laboratory, and electrocardiography (ECG) parameters. In our included studies, tenapanor therapy didn’t lead to any significant change in physical examination, laboratory, and ECG parameters. There was also no death related to tenapanor. From these findings, tenapanor was relatively safe and well tolerated in CKD patients.^7–9,11,12^

## Discussion

Phosphate retention is frequently found in CKD stage 4 and 5, which is a factor that initiate many other disturbances such as increased FGF23 and PTH, hypocalcemia, and low vitamin D, which in turn will lead to an enhancement in cardiovascular and all cause of morbidity and mortality.^16^ Phosphate balance is maintained by several mechanism including intestinal phosphate absorption, bone turnover regulation, as well as renal excretion and reabsorption.^4,17^ In a normal condition, kidney excretes approximately ninety percent of phosphate per day.^18^ As renal phosphate clearance declined in CKD, inhibiting intestinal phosphate absorption can be a promising approach in managing hyperphosphatemia in patients with CKD.^4,18^

Generally, phosphate binders only result in maximum 2.0 mg/dL of phosphate reduction in their maximal dose.^7^ Some of our included studies show that 30 mg tenapanor whether as fixed or titrated dose single therapy was able to reduce serum phosphate up to more than 2.0 mg/dL.^7,8^ This finding can be possible due to tenapanor mechanism which suppress intestinal phosphate absorption through NHE3 inhibition^7–9,12^, as about 90% of phosphate input comes from intestinal absoption.^18^

In addition to its phosphate lowering effect, tenapanor was also found to have a pleiotropic effect in suppressing the levels of FGF23 and PTH, both as monotherapy and combination therapy with phosphate binders.^8–11,13,14^ The exact mechanism of this finding remains unclear, but it can be possibly explained by the role of these two hormones as compensatory mechanism to elevated serum phosphate, thus greater reduction of serum phosphate will suppress FGF23 and PTH production.^7,9,10,13^ This pleiotropic effect of tenapanor can be utilized to minimize the morbidity and mortality in CKD, as high FGF23 and PTH will cause extraosseous calcium and phosphate deposition, particularly in the heart valves and vascular. This condition then develop to valve and vascular atherosclerosis through osteochondrogenic differentiation and elastin degradation, leading to increased mortality due to cardiovascular disease.^19^ Besides that, increased PTH which enhance calcium efflux from bones is the cause of bone mass loss in patients with CKD, resulting in increased morbidity.^20^ Therefore, the pleiotropic effect of tenapanor in lowering FGF23 and PTH suggesting its potency as future treatment for bone mineral disease and secondary hyperparathyroidism in CKD.

Diarrhea is the most common AE caused by tenapanor. However, majority of reported diarrhea was transient and mild to moderate in intensity.^11–13^ This side effect appears as the effect of tenapanor mechanism which selectively inhibit the NHE3 in enterocytes, thus suppress the passive transport of phosphate in the intestine. Simultaneously, NHE3 inhibition also suppress intestinal sodium absorption which enhance sodium and water secretion in the intestinal tract.^8,9^ Elevated sodium, phosphate, and water content in the intestine then lead to loose stool and increased bowel movement which manifested as diarrhea.^7,13^ Incidence of diarrhea due to tenapanor has a higher trend in higher dose.^7–13^ Dose titration can be a good solution for this issue to increase patient’s compliance, as titrated dose of tenapanor 30 mg BID exhibited a lower incidence of diarrhea while maintaining the same efficacy in reducing serum phosphate and FGF23 compared to fixed dose of tenapanor 30 mg BID.^9^

Minerals deposition and metabolic disturbances are the other concerns in treatment options for hyperphosphatemia in CKD. Available phosphate binders still has systemic mineral and metabolic disadvantages, such as calcium accumulation and increased calcification in calcium based phosphate binders, potential nervous system toxicity in aluminium based phosphate binders, iron overload in iron based phosphate binders, fat-soluble vitamins deficiency and acidosis risk in sevelamer based phosphate binders, as well as possible lanthanum deposits in lanthanum based phosphate binders.^4–6^ Tenapanor comes as a solution to counteract these limitations, as tenapanor didn’t cause any meaningful changes in serum calcium, laboratory parameters, ECG, and physical examination.^7–9,11,12^ The plausible explanation for this finding is the nature of tenapanor as a calcium free non-metal phosphate reducer, thus it doesn’t cause calcium and metal accumulation. In addition, tenapanor is also a minimally absorbed molecule, resulting in minimum systemic side-effect.^7,8^

## CONCLUSION AND RECOMMENDATION

In conclusion, tenapanor, as single therapy or combination with phosphate binders, was effective in managing hyperphosphatemia in CKD. It also has pleiotropic effects in interfering high FGF23 and PTH thus exhibit a promising potency to treat secondary hyperparathyroidism in CKD. Diarrhea is the most common AE but it’s only transient, dose-related, and mild to moderate in severity. Starting tenapanor with low dose can be a solution for diarrhea side effect. Tenapanor doesn’t cause any meaningful change in ECG, serum calcium, and other laboratory and clinical parameters. Generally, tenapanor is safe and well-tolerated in CKD patients.

## Data Availability

All data produced in the present study are available upon reasonable request to the authors
All data produced in the present work are contained in the manuscript

## ACKNOWLEDGMENT

-

